# Life-course Stress Exposure and Cognitive Decline in Middle-aged and Older Chinese Adults: The Role of Sex Differences and Educational Protection

**DOI:** 10.64898/2026.01.26.26344833

**Authors:** Jiahao Li, Giulia Mozzanica, Feng Zhang, Jocelien D. A. Olivier, Ulrich L. M. Eisel

## Abstract

**Background:** Stressful life events (SLEs) across the life course have been associated with cognitive decline, but evidence on their cumulative impact and potential modifiers remains limited. We aimed to examine the associations between SLE exposure in childhood, adulthood, or both life stages and cognitive trajectories, and to investigate whether these associations vary by sex and level of education.

**Methods:** We used data from the China Health and Retirement Longitudinal Study (CHARLS), a nationally representative cohort of adults aged 45 years and older. Participants with complete data on SLEs, cognitive function, and covariates were included (n=5,922). SLEs were assessed retrospectively for childhood and adulthood periods. Cognitive function was measured using a composite score (range 0–21) across three waves (2011–2015). Linear mixed-effects models were used to examine longitudinal associations, adjusting for sociodemographic factors, health behaviors, and chronic health conditions. Interaction analyses were performed to explore effect modification by sex and education.

**Results:** Of 5,922 participants, 2,428 (41.0%) reported no SLEs, 1,374 (23.2%) experienced childhood-only SLEs, 1,294 (21.8%) experienced adulthood-only SLEs, and 826 (14.0%) had cumulative SLE exposure (both childhood and adulthood). In fully adjusted longitudinal models, cumulative SLE exposure showed the strongest association with cognitive decline (β = –0.52, 95% CI –0.69 to –0.35, p<0.001), followed by childhood-only (β = –0.34, –0.48 to –0.20, p<0.001) and adulthood-only exposure (β = –0.22, –0.37 to –0.07, p<0.01). Sex significantly moderated the associations for childhood-only and cumulative exposure (interaction β = –0.21, 95% CI –0.36 to –0.05, p<0.01; interaction β = –0.25, 95% CI –0.43 to –0.07, p<0.01), with women showing greater cognitive vulnerability than men. Higher educational attainment buffered against single-period stress effects but provided only partial protection against cumulative life-course adversity.

**Conclusion:** Life-course stress exposure, particularly when cumulative, is associated with accelerated cognitive decline in Chinese middle-aged and older adults. Women show greater vulnerability to stress-related cognitive effects, whereas higher education appears to buffer against such impacts across both sexes. These findings underscore the importance of sex-sensitive, education-informed approaches to stress reduction and cognitive health promotion across the life course.

## 1. Introduction

Cognitive decline represents one of the most significant challenges in global population ageing, with profound implications for individual wellbeing, healthcare systems, and societal functioning(1). While age remains the strongest risk factor for cognitive impairment, mounting evidence suggests that psychosocial stressors across the life course substantially influence cognition, potentially accounting for considerable heterogeneity in cognitive ageing patterns(2, 3).

Stressful life events (SLEs) encompass major adverse experiences such as bereavement, financial hardship, serious illness, and interpersonal trauma that disrupt individuals’ psychological equilibrium and adaptive capacity(4, 5). The neurobiological consequences of stress exposure are well-documented: chronic activation of the hypothalamic-pituitary-adrenal axis leads to elevated glucocorticoid levels(6), which can damage hippocampal neurons, impair synaptic plasticity, and accelerate brain ageing(7, 8). Additionally, stress-induced inflammatory processes and oxidative stress contribute to neurodegeneration and cognitive dysfunction(9). However, the timing of stress exposure may differentially affect cognitive outcomes through distinct mechanisms.

Childhood represents a critical neurodevelopmental period when the brain exhibits heightened plasticity but also vulnerability to environmental insults(10, 11). Early-life adversity can result in lasting alterations to stress-response systems, brain structure, and cognitive reserve that persist into later life(12, 13). In contrast, stress exposure in adulthood may deplete existing cognitive resources and accelerate age-related neurodegeneration through allostatic load(14). Accordingly, stressful life events occurring in childhood and adulthood are conceptualized as distinct exposures and assessed separately (see Methods for details). Despite this theoretical framework, few studies have simultaneously examined the independent and cumulative effects of childhood and adulthood SLEs on cognitive function using a life-course approach.

Furthermore, individual vulnerability to stress-related cognitive effects likely varies across demographic groups. Sex differences in stress reactivity, coping strategies, and neurobiological responses have been observed(15), with some evidence suggesting women may show a faster rate of cognitive decline(16). Educational attainment, a key component of cognitive reserve, may confer protection against cognitive decline through enhanced compensatory mechanisms and neural efficiency(17, 18). However, whether and how sex and education modify the relationship between life-course stress exposure and cognition remains largely unexplored. Most existing studies have examined these factors in isolation rather than as potential effect modifiers. Moreover, understanding these relationships within specific cultural and socioeconomic contexts is crucial, as stress experiences, gender roles, and educational systems are inherently shaped by local cultural frameworks and historical trajectories(19–21).

In this study, we investigated associations between life-course patterns of stress exposure and cognitive function in a large Chinese cohort of middle-aged and older adults. We examined whether the timing of stress exposure—during childhood, adulthood, or both periods—relates differentially to cognitive outcomes, and assessed the extent to which these associations vary by sex and educational attainment. Evidence from this population-based cohort study could help clarify the contribution of psychosocial stress to cognitive ageing and inform approaches to risk stratification and prevention.

## 2. Material and methods

### 2.1 Study design and participants

This study used data from the China Health and Retirement Longitudinal Study (CHARLS), a nationally representative longitudinal survey of adults aged 45 years and older. CHARLS employed a multistage stratified probability-proportionate-to-size sampling design, covering 450 villages and communities across 28 provinces in mainland China(22). The baseline survey was conducted in 2011, with follow-up waves in 2013 and 2015. The 2014 CHARLS Life History Survey provided retrospective information on childhood experiences.

For this analysis, we merged data from waves 1–3 (2011–2015) with the 2014 Life History Survey. Participants were included if they: (1) completed both the Life History Survey and main surveys; (2) were aged 45 years or older at baseline; (3) had complete data on SLEs and cognitive assessments; and (4) had no missing data on key covariates. Participants previously diagnosed with dementia or Alzheimer’s disease were excluded, yielding a final analytic sample of 5,922 participants. The study was approved by the Institutional Review Board of Peking University (IRB00001052-11015). All participants provided written informed consent.

### 2.2 Assessment of stressful life events

SLEs were assessed using validated items from the CHARLS Life History Survey for childhood events and from the baseline survey (2011) for adulthood events, and each item was dichotomized (yes/no) based on established criteria (Supplementary Table 1). Childhood SLEs included: financial hardship, parental unemployment, parental substance abuse, foster care placement, parental separation or divorce, and parental death. Adulthood SLEs included: unemployment, asset poverty, death of a child, death of a spouse, life-threatening illness or accident, and physical assault. The selection of SLE indicators was based on the criteria described by Zhou et al.(3), and participants were categorized into four mutually exclusive groups: (1) no SLEs (reference group); (2) childhood SLEs only; (3) adulthood SLEs only; and (4) both childhood and adulthood SLEs.

### 2.3 Cognitive assessment

Cognitive function was measured using a standardized battery adapted from the U.S. Health and Retirement Study, with components analyzed both globally and by domain(23). Memory performance (range 0–10) was assessed as the average of immediate and delayed recall of ten words. Executive function (range 0–6) was assessed using one item from the Telephone Interview for Cognitive Status, 10-item version (TICS-10; serial 7 subtraction, performed five times), together with a figure-drawing task that also captured visuospatial ability. Orientation (range 0–5) was evaluated using five temporal items from the TICS-10, including current year, month, date, day of week, and season. Global cognitive function (range 0–21) was calculated as the sum of episodic memory, executive function, and orientation scores. Higher values indicated better performance in each domain. This cognitive battery has been widely used in international ageing studies and has demonstrated good reliability and validity, including in Chinese populations(24).

### 2.4 Covariates

Key covariates were selected based on established risk factors for cognitive decline(1). Sociodemographic variables included age (continuous and categorized as <60 or ≥60 years), sex, educational attainment (below high school vs high school or above), and residence (rural vs urban). Health behaviors included smoking status (never vs current/former smokers) and alcohol consumption (<1 drink/week vs ≥1 drink/week). Health-related variables included body mass index (normal range 18.5–23.9 kg/m² vs others) and physician-diagnosed chronic health conditions (hypertension, diabetes, and cardiovascular disease).

### 2.5 Statistical and Sensitivity Analyses

We described baseline characteristics by SLE exposure group using means and standard deviations for continuous variables and frequencies and percentages for categorical variables. We examined associations between SLE exposure and global cognitive function and domain-specific cognitive performance (memory, executive function, orientation). Cross-sectional associations between SLE exposure and baseline cognitive scores were examined using ordinary least squares regression. Longitudinal associations were assessed using linear mixed-effects models with random intercepts to account for within-person correlation across waves. Two models with increasing adjustment were fitted: Model 1 was unadjusted; Model 2 adjusted for age, sex, education, residence, health behaviors and chronic conditions.

In addition, subgroup analyses were performed based on key covariates to explore potential heterogeneity in the associations between SLE exposure and cognitive outcomes. To investigate effect modification, we tested interaction terms between SLE exposure and age, sex, and education— selected for their theoretical importance as determinants of cognitive reserve and stress vulnerability(1). Bootstrap resampling (1000 iterations) was used to ensure robust inference for these interaction tests. Based on the interaction analyses, we conducted stratified analyses by sex and education to examine group-specific associations. Sensitivity analyses compared random intercept versus random slope models to assess robustness of findings. All analyses were performed using R version 4.4.3. Statistical significance was set at two-sided p<0.05.

## 3. Results

### 3.1 Participant characteristics

Of 5,922 participants included in the analysis, 2,428 (41.0%) reported no SLEs, 1,374 (23.2%) experienced childhood-only SLEs, 1,294 (21.8%) experienced adulthood-only SLEs, and 826 (14.0%) reported SLEs in both life stages (Table 1). The mean age was 56.5 years (SD 8.5). The mean age for males (3,212; 54.2%) was 57.7 ± 8.4 years, while the mean age for women (2,710; 45.8%) was 55.0 ± 8.5 years. Of all participants 4,958 (83.7%) had below high school education.

**Table 1.** Baseline characteristics of participants according to stressful life event exposure group.

| Characteristics | Total | None | Childhood only | Adulthood only | Childhood and adulthood |
| --- | --- | --- | --- | --- | --- |
| **Reports** (n, %) | 5,922 (100%) | 2,428 (41.0%) | 1,374 (23.2%) | 1,294 (21.8%) | 826 (14.0%) |
| **Age** (n, %) |  |  |  |  |  |
| <60 | 3,780 (63.8%) | 1,660 (68.4%) | 835 (60.8%) | 816 (63.1%) | 469 (56.8%) |
| ≥60 | 2,142 (36.2%) | 768 (31.6%) | 539 (39.2%) | 478 (36.9%) | 357 (43.2%) |
| **Sex** (n, %) |  |  |  |  |  |
| Male | 3,212 (54.2%) | 1,267 (52.2%) | 784 (57.1%) | 672 (51.9%) | 489 (59.2%) |
| Female | 2,710 (45.8%) | 1,161 (47.8%) | 590 (42.9%) | 622 (48.1%) | 337 (40.8%) |
| **Educational attainment** (n, %) |  |  |  |  |  |
| Below high school | 4,958 (83.7%) | 1,933 (79.6%) | 1,187 (86.4%) | 1,098 (84.9%) | 740 (89.6%) |
| High school or above | 964 (16.3%) | 495 (20.4%) | 187 (13.6%) | 196 (15.1%) | 86 (10.4%) |
| **Residence** (n, %) |  |  |  |  |  |
| Urban | 2,478 (41.8%) | 1,135 (46.7%) | 525 (38.2%) | 522 (40.3%) | 296 (35.8%) |
| Rural | 3,444 (58.2%) | 1,293 (53.3%) | 849 (61.8%) | 772 (59.7%) | 530 (64.2%) |
| **Smoke** (n, %) |  |  |  |  |  |
| Never smokers | 3,382 (57.1%) | 1,477 (60.8%) | 751 (54.7%) | 741 (57.3%) | 413 (50.0%) |
| Current or former smokers | 2,540 (42.9%) | 951 (39.2%) | 623 (45.3%) | 553 (42.7%) | 413 (50.0%) |
| **Alcohol consumption** (n, %) |  |  |  |  |  |
| <1 drink/week | 3,390 (57.2%) | 1,436 (59.1%) | 760 (55.3%) | 750 (58.0%) | 444 (53.8%) |
| ≥1 drink/week | 2,532 (42.8%) | 992 (40.9%) | 614 (44.7%) | 544 (42.0%) | 382 (46.2%) |
| **BMI category (n, %)** |  |  |  |  |  |
| Normal range | 3,199 (54.0%) | 1265 (52.1%) | 731 (53.2%) | 719 (55.6%) | 484 (58.6%) |
| Others | 2,723 (46.0%) | 1163 (47.9%) | 643 (46.8%) | 575 (44.4%) | 342 (41.4%) |
| **Hypertension (n, %)** |  |  |  |  |  |
| Yes | 1,497 (25.3%) | 607 (25.0%) | 352 (25.6%) | 322 (24.9%) | 216 (26.2%) |
| No | 4,425 (74.7%) | 1,821 (75.0%) | 1,022 (74.4%) | 972 (75.1%) | 610 (73.8%) |
| **Diabetes (n, %)** |  |  |  |  |  |
| Yes | 378 (6.4%) | 161 (6.6%) | 79 (5.7%) | 95 (7.3%) | 43 (5.2%) |
| No | 5,544 (93.6%) | 2,267 (93.4%) | 1,295 (94.3%) | 1,199 (92.7%) | 783 (94.8%) |
| **Cardiovascular disease (n, %)** |  |  |  |  |  |
| Yes | 741 (12.5%) | 277 (11.4%) | 183 (13.3%) | 165 (12.8%) | 116 (14.0%) |
| No | 5,181 (87.5%) | 2,151 (88.6%) | 1,191 (86.7%) | 1,129 (87.2%) | 710 (86.0%) |
| **Global cognitive score (mean ± SD)** | 13.2 ± 2.8 | 13.5 ± 2.6 | 13.0 ± 2.8 | 13.1 ± 2.8 | 12.7 ± 2.8 |

Values are presented as mean ± standard deviation (SD) for continuous variables and number (percentage) for categorical variables.

Participants with cumulative SLE exposure (both childhood and adulthood) differed from other groups in several characteristics. Although males outnumbered females in the overall study population, the cumulative SLE exposure group showed the largest male–female imbalance (489 [59.2%] males vs 337 [40.8%] females) compared with the other exposure groups. They were also more likely to reside in rural areas (530 [64.2%] vs 1,293 [53.3%]), have lower educational attainment (740 [89.6%] vs 1,933 [79.6%]), and report current smoking (413 [50.0%] vs 951 [39.2%]) (additional characteristics are presented in Table 1). Mean baseline cognitive scores decreased with increasing SLE exposure: 13.5 (SD 2.6) for no SLEs, 13.0 (2.8) for childhood-only, 13.1 (2.8) for adulthood-only, and 12.7 (2.8) for both childhood and adulthood SLEs.

### 3.2 Cross-sectional associations

In unadjusted analyses (Model 1), all SLE exposure groups showed significantly lower baseline cognitive scores compared with the reference group (Table 2). The largest difference was observed for cumulative exposure (β = –0.76, 95% CI –0.97 to –0.54, p<0.001), followed by childhood-only (β = –0.45, 95% CI –0.63 to –0.26, p<0.001) and adulthood-only exposure (β = –0.39, 95% CI –0.58 to –0.21, p<0.001). After full adjustment for sociodemographic factors, health behaviors, and chronic conditions (Model 2), associations remained significant for childhood-only (β = –0.25, –0.43 to –0.08, p<0.01) and cumulative exposure (β = –0.41, –0.62 to –0.19, p<0.001), while adulthood-only exposure showed a non-significant trend (β = –0.17, – 0.35 to 0.01, p=0.071).

**Table 2.** Cross-sectional and longitudinal associations between stressful life events and global cognitive function across adjustment models.

| Model | None | Childhood only | Adulthood only | Childhood and adulthood |
| --- | --- | --- | --- | --- |
| Value | Ref | $\beta$ (95% CI) | $\beta$ (95% CI) | $\beta$ (95% CI) |
| *Cross-sectional* |  |  |  |  |
| **Model 1** | Ref | -0.45 (-0.63, -0.26)<br>$p < 0.001^{***}$ | -0.39 (-0.58, -0.21)<br>$p < 0.001^{***}$ | -0.76 (-0.97, -0.54)<br>$p < 0.001^{***}$ |
| **Model 2** | Ref | -0.25 (-0.43, -0.08)<br>$p < 0.01^{**}$ | -0.17 (-0.35, 0.01)<br>$p = 0.071$ | -0.41 (-0.62, -0.19)<br>$p < 0.001^{***}$ |
| *Longitudinal* |  |  |  |  |
| **Model 1** | Ref | -0.56 (-0.71, -0.40)<br>$p < 0.001^{***}$ | -0.49 (-0.64, -0.33)<br>$p < 0.001^{***}$ | -0.93 (-1.12, -0.75)<br>$p < 0.001^{***}$ |
| **Model 2** | Ref | -0.34 (-0.48, -0.20)<br>$p < 0.001^{***}$ | -0.22 (-0.37, -0.07)<br>$p < 0.01^{**}$ | -0.52 (-0.69, -0.35)<br>$p < 0.001^{***}$ |

Notes: Values represent regression coefficients ( $\beta$ ), 95% confidence intervals, and p-values for the association between SLE exposure and global cognitive function.
The reference group is individuals without any reported SLE exposure (“None”).
Model 1 was unadjusted.
Model 2 additionally adjusts for age, sex, education, residence, smoking and drinking status, BMI category, hypertension, diabetes, and cardiovascular disease.
$^{***} p < 0.001$ , $^{**} p < 0.01$ , $^{*} p < 0.05$

### 3.3 Longitudinal cognitive trajectories

Over the 4-year follow-up period, cognitive function declined in all groups, with steeper trajectories observed among those with SLE exposure (Figure 1). The direction of associations remained consistent between unadjusted and fully adjusted models (Table 2). In fully adjusted longitudinal models (Model 2), all SLE exposure groups showed significant associations with cognitive decline compared with the no-SLE group. The strongest association was observed for cumulative exposure (β = – 0.52, 95% CI –0.69 to –0.35, p<0.001), followed by childhood-only (β = –0.34, –0.48 to –0.20, p<0.001) and adulthood-only exposure (β = –0.22, –0.37 to –0.07, p<0.01).

**Figure 1.**
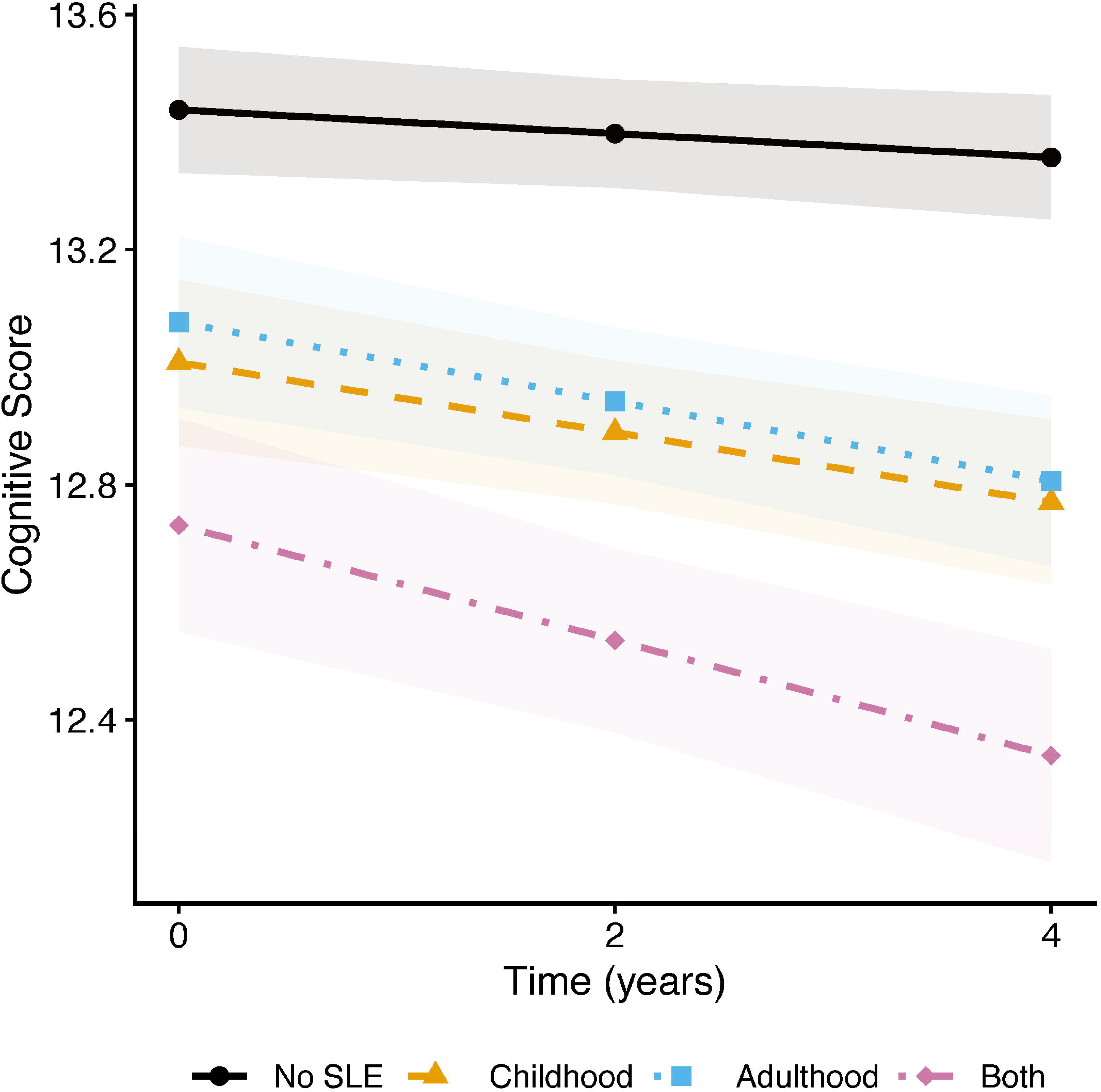
Global cognition trajectories across SLE exposure groups. Longitudinal cognitive trajectories over 4 years in Chinese middle-aged and older adults with 95% confidence intervals. Groups: no SLE (black), childhood only (orange), adulthood only (blue), and both periods (purple). Cumulative exposure shows the steepest decline.

Domain-specific analyses of memory, executive function, and orientation revealed differential patterns by timing of stress exposure (Supplementary Tables 2–4). In fully adjusted longitudinal models, childhood-only exposure showed stronger associations with memory decline (β = –0.18, 95% CI –0.26 to –0.10, p<0.001) than with executive function (β = –0.02, –0.08 to 0.05, p=0.662) or orientation (β = –0.02, –0.06 to 0.02, p=0.331). Conversely, adulthood-only exposure demonstrated consistent associations with executive function across both baseline and longitudinal analyses, whereas its associations with memory and orientation were not significant at baseline but became significant in longitudinal models (memory: β = –0.12, –0.20 to –0.04, p<0.01; orientation: β = –0.07, –0.11 to –0.03, p<0.01). Cumulative exposure showed significant associations across all three domains, with the strongest effect on memory (β = –0.26, –0.35 to –0.16, p<0.001).

### 3.4 Subgroup and sensitivity analyses

In the subgroup analyses, several sociodemographic and health-related factors were independently associated with poorer cognitive trajectories (Table 3). Older age (≥60 years: β = –0.54, 95% CI –0.63 to –0.44, p<0.001), female (β = –0.61, –0.76 to –0.45, p<0.001), and lower educational attainment (below high school: β = –1.64, – 1.79 to –1.48, p<0.001) showed strong associations with cognitive decline. In addition, place of residence was an important determinant of cognitive outcomes. Those living in rural areas (β = –0.76, 95% CI –0.88 to –0.64, p<0.001) had poorer cognitive trajectories compared with their urban counterparts. Among health-related and behavioral factors, current or former smoking (β = –0.19, 95% CI –0.33 to –0.05, p<0.01) and hypertension (β = –0.19, 95% CI –0.30 to –0.08, p<0.001) were both independently associated with faster cognitive decline. Sensitivity analyses revealed highly consistent estimates between the random slope and random intercept models (Table 3), with effect sizes and significance levels remaining stable for all SLE exposure groups and covariates, confirming the robustness of the findings.

**Table 3.**
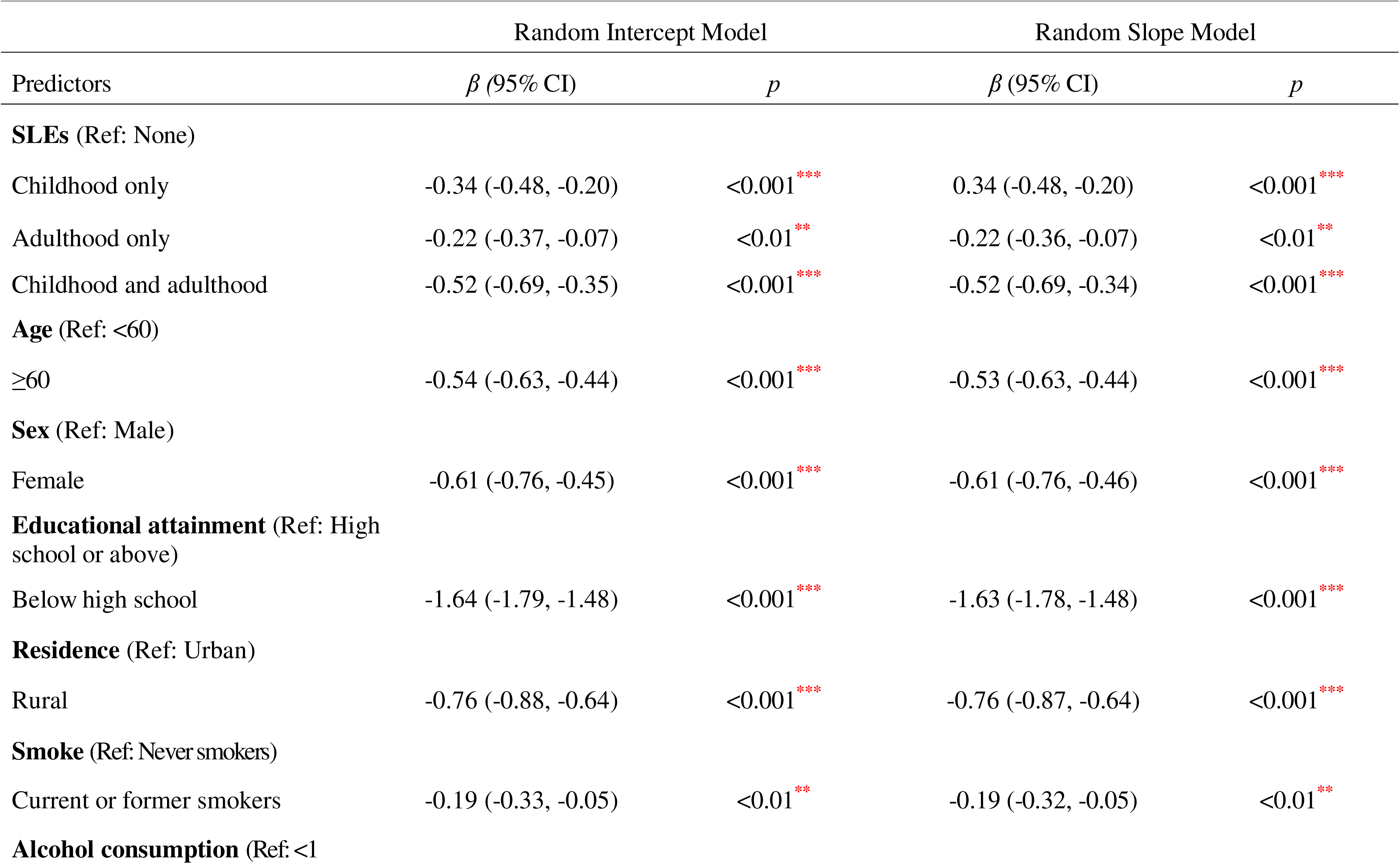
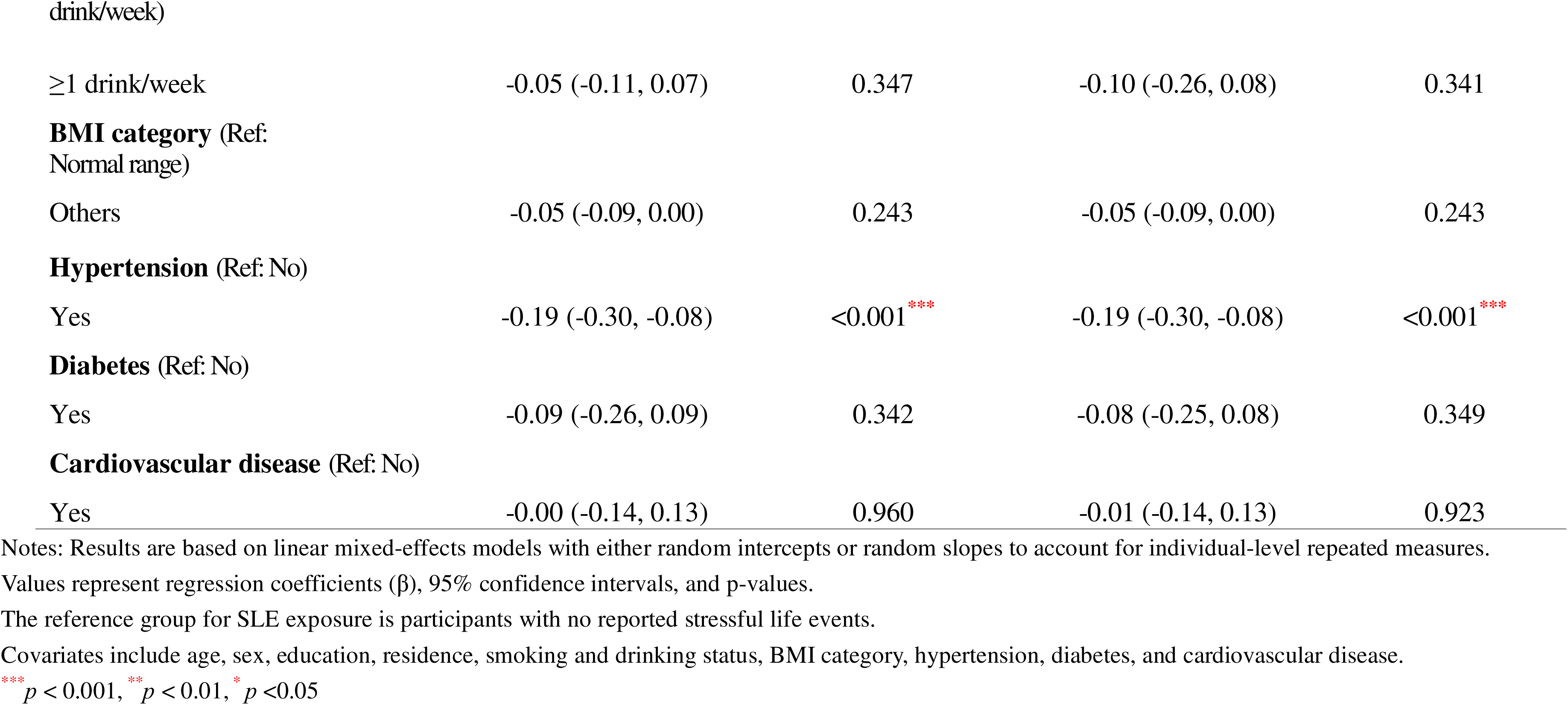
Comparison of random intercept and random slope models for the association between stressful life events and global cognitive fun<u>ction</u>.

### 3.5 Effect modification

Interaction analyses revealed significant effect modification by sex on cognitive outcomes for both childhood-only (interaction β = –0.21, 95% CI –0.36 to – 0.05, p<0.01) and cumulative SLE exposure (interaction β = –0.25, 95% CI –0.43 to – 0.07, p<0.01), but not for adulthood-only exposure (Figure 2A). Sex-stratified analyses confirmed differential vulnerability: among women, all SLE exposure groups showed stronger associations than in men (Figure 2B). For cumulative exposure, women showed a –1.02-point difference (95% CI –1.33 to –0.71, p<0.001) compared with –0.79 (–1.00 to –0.58, p<0.001) among men in fully adjusted models.

**Figure 2.**
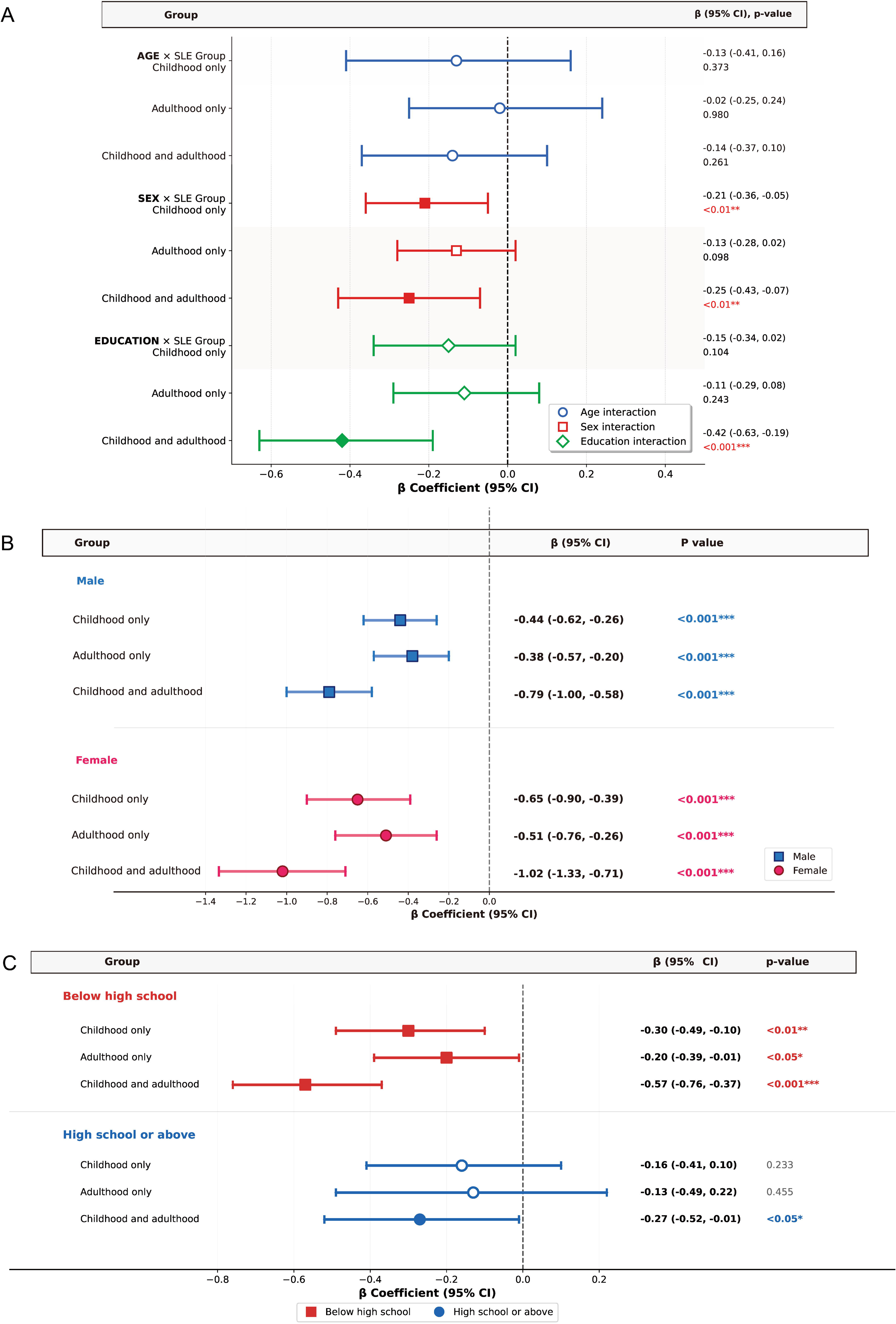
Interaction and stratified associations between SLE exposure and cognitive function. **(A)** Forest plot of interaction effects between SLE exposure and demographic factors. Interaction coefficients with 95% confidence intervals for age (blue circles), sex (red squares), and education (green diamonds) with SLE exposure groups. Filled symbols indicate significance. Significant interactions observed for sex and education with cumulative SLE exposure. **(B)** Sex-stratified associations between SLE exposure and cognitive function. Forest plot showing associations for males (blue squares) and females (red circles). Reference: no SLE. Women show consistently stronger associations across all SLE patterns, particularly for cumulative exposure. **(C)** Education-stratified associations between SLE exposure and cognitive function. Forest plot showing associations for below high school (red squares) and high school or above (blue circles). Filled symbols indicate significance. Higher education demonstrates a protective buffering effect against stress-related cognitive impacts. ***p<0.001, **p<0.01, *p<0.05.

Education also showed significant interaction effects with cumulative SLE exposure on cognitive outcomes (interaction β = –0.42, 95% CI –0.63 to –0.19, p<0.001), but not with childhood-only or adulthood-only exposure (Figure 2A). Education-stratified analyses indicated a strong protective effect of higher educational attainment. Among participants with less than high school education, all SLE exposure groups were significantly associated with cognitive decline in fully adjusted models (Figure 2C), including cumulative exposure (β = –0.57, 95% CI –0.76 to – 0.37, p < 0.001), childhood-only exposure (β = –0.30, 95% CI –0.49 to –0.10, p < 0.01), and adulthood-only exposure (β = –0.20, 95% CI –0.39 to –0.01, p < 0.05).

In contrast, among participants with high school education or above, these associations were substantially attenuated, and cumulative exposure remained significant (β = –0.27, 95% CI: –0.52 to –0.01, p < 0.05), while childhood-only and adulthood-only exposures were no longer significant (Figure 2C). Additional details on interaction effects and stratified analyses are provided in Supplementary Tables 5–7.

## 4. Discussion

In this large longitudinal cohort of Chinese middle-aged and older adults, we found that exposure to SLEs across the life course was significantly associated with poorer cognitive function and steeper cognitive decline over time. The association was strongest for individuals exposed to stressors in both childhood and adulthood, supporting a cumulative stress hypothesis(6). Notably, women showed greater vulnerability to stress-related cognitive effects, while higher educational attainment appeared to confer substantial protection. These findings provide important evidence for life-course approaches to understanding cognitive ageing in non-Western populations.

Our results align with emerging literature suggesting that psychosocial stress accumulates across the life span to influence brain health(6, 25–27). The dose-response relationship observed supports allostatic load theory, whereby cumulative stress exposure across different life stages overwhelms adaptive capacity and accelerates neurodegeneration(28). The persistent associations after adjusting for multiple health and lifestyle factors suggest that stress effects on cognition operate through pathways beyond traditional cardiovascular and metabolic risk factors, potentially involving direct neurotoxic effects of glucocorticoid exposure, stress-induced neuroinflammation, and disruption of neurotrophic support systems(14).

Recent evidence suggests that the cumulative effects of life-course stress may involve stress sensitization mechanisms(29). Specifically, childhood adversity can induce lasting epigenetic and neurobiological alterations that sensitize individuals to future stressors(30, 31). This biological embedding process creates a vulnerability that amplifies the impact of adult stress on cognitive function(32). Such programming of stress-response systems during sensitive developmental periods may fundamentally alter trajectories of cognitive aging, providing a mechanistic framework for understanding why individuals with both childhood and adult stress exposure showed the poorest cognitive outcomes in our study.

Our domain-specific analyses provide novel insights into the neurocognitive signatures of stress exposure timing(6). The selective vulnerability of memory systems to childhood stress aligns with extensive evidence that early-life adversity specifically disrupts hippocampal development during critical periods of postnatal neurogenesis and synaptic pruning(33, 34). The hippocampus, with its high density of glucocorticoid receptors and protracted development extending into adolescence, appears particularly susceptible to early stress exposure(35). Conversely, the broader cognitive impact of adulthood stress was characterized by differential domain vulnerability: executive function showed both immediate and progressive impairments, whereas memory and orientation declined primarily over time. This pattern may reflect how stress affects mature neural systems already undergoing age-related changes(6).

The implications for intervention design are substantial. Memory-focused cognitive training may be particularly beneficial for individuals with childhood trauma histories, potentially helping to compensate for early hippocampal disruption. For those experiencing adult-life stressors, comprehensive interventions combining stress management with broader cognitive stimulation targeting executive networks may prove more effective. The finding that cumulative exposure produced the strongest effects across all domains underscores the importance of prevention strategies that span the entire life course rather than focusing on single developmental periods.

Sex differences in stress vulnerability likely reflect complex interplay between biological and social factors. Women’s greater susceptibility may stem from sex differences in stress neurobiology, including enhanced HPA axis reactivity(36), greater inflammatory responses to psychosocial stressors(37), and hormonal fluctuations that modulate stress sensitivity across the lifespan(38). Additionally, sex-specific social experiences—including differential exposure to interpersonal stressors, caregiving burdens, and economic disadvantages—may amplify vulnerability to stress-related cognitive decline(39, 40). The absence of sex interaction for adulthood-only stress suggests these vulnerabilities may be particularly pronounced when stress occurs early in development or accumulates over time, potentially reflecting organizational effects of early stress on developing stress-response systems(41). However, sex differences in stress-related cognitive outcomes may not follow a uniform pattern. In our previous work, poorer cognitive performance was observed among men in a study context involving multiple stressors in adulthood, suggesting that male vulnerability may vary under conditions of high stress load(4). Such discrepancies may reflect differences in stress characteristics, cumulative burden, and population context, underscoring the context-dependent nature of sex differences in stress-related cognitive decline.

The protective effect of education likely operates through multiple interconnected pathways. Beyond providing cognitive reserve through enhanced neural efficiency and compensatory networks(42), education shapes lifelong trajectories of health behaviors, occupational complexity, and social capital that buffer against stress(43, 44). The near-complete attenuation of single-period stress effects among educated individuals suggests cognitive reserve may be particularly effective against isolated stressors. However, even education cannot fully compensate for cumulative life-course adversity, indicating limits to cognitive reserve when allostatic load exceeds compensatory capacity. This finding has important implications for societies with limited educational access, where cognitive vulnerability to stress may be amplified at the population level(45).

The independent effects of rural residence highlight how stress operates within broader contexts of social disadvantage. Rural-urban disparities in cognitive outcomes likely reflect complex interactions between healthcare access, cognitive stimulation opportunities, environmental exposures, and social network characteristics(46). Social connections and supportive environments may provide protection through multiple mechanisms, including emotional support, cognitive engagement, and shared resources that buffer against stress(47). Our findings underscore that addressing stress-related cognitive decline requires attention to broader social determinants of brain health.

Our findings have important public health implications for ageing societies globally. The identification of stress as a modifiable risk factor for cognitive decline suggests potential for prevention through stress-reduction interventions implemented across the life course. Early childhood programs addressing adverse experiences could prevent the establishment of neurobiological vulnerability, while workplace stress management and community-based resilience programs in adulthood might slow cognitive ageing trajectories. The particular vulnerability of women suggests the need for sex-sensitive approaches that address both biological and social sources of differential risk.

Several limitations warrant consideration. Retrospective assessment of SLEs may introduce recall bias, though major life events are typically well-remembered and any misclassification would likely attenuate observed associations. The inability to assess stress chronicity, severity, or subjective appraisal limits understanding of dose-response relationships. While our longitudinal design strengthens causal inference, reverse causality cannot be definitively excluded. While our 4-year follow-up period aligns with established practices in cognitive research(48, 49), extended longitudinal observation would enhance our understanding of stress-related cognitive trajectories. Finally, cultural variations in stress perception, expression, and coping may limit generalizability to other populations, though the biological mechanisms linking stress to cognitive decline are likely universal.

Future research should investigate neurobiological mechanisms through neuroimaging studies examining stress-related changes in brain structure and function across the lifespan. Intervention trials testing targeted stress-reduction strategies at different life stages would provide causal evidence for prevention potential. Examining protective factors beyond education—including specific coping strategies, social support quality, and lifestyle factors—could identify additional intervention targets. Integration of biomarkers of stress exposure and allostatic load would strengthen understanding of biological pathways linking psychosocial stress to cognitive outcomes.

## 5. Conclusion

In conclusion, cumulative stress exposure across the life course is associated with accelerated cognitive decline, with particular vulnerability among women and protection conferred by education. These findings underscore the importance of conceptualizing cognitive health within a life-course framework that recognizes the enduring impact of psychosocial adversity. Public health strategies incorporating early intervention for childhood adversity, stress management across adulthood, sex-sensitive approaches, and expanded educational opportunities may help preserve cognitive function in ageing populations worldwide.

## Acknowledgements

Not applicable.

## CRediT authorship contribution statement

Jiahao Li: Conceptualization; Methodology; Data curation; Formal analysis; Writing – original draft; Writing – review and editing. Giulia Mozzanica: Writing – review and editing; Support with literature review and manuscript revision. Feng Zhang: Conceptualization; Supervision; Writing – review and editing. Jocelien D. A. Olivier: Supervision; Writing – review and editing; Critical revision of the manuscript. Ulrich L. M. Eisel: Conceptualization; Project administration; Supervision; Writing – review and editing; Critical revision of the manuscript.

## Funding

This work was supported by funding from Alzheimer Nederland (WE.03-2024-20, Ulrich L. M. Eisel and Jocelien D. A. Olivier) and the National Natural Science Foundation of China (NSFC 82401393, Feng Zhang).

## Declaration of competing interest

The authors declare that they have no competing interests.

## Ethical approval

All participants provided written informed consent, and ethical approval was obtained from the Institutional Review Board of Peking University (IRB00001052–11015)

## Data Availability

The datasets analyzed during the current study are available from the China Health and Retirement Longitudinal Study repository (https://charls.pku.edu.cn/en/) upon reasonable request and application to the data custodians. All data generated or analyzed in this study are included in this published article.

## Declaration of Generative AI and AI-assisted technologies in the writing process

No generative AI and AI-assisted technologies were used in the writing process of this work.

## Notes

### Competing Interest Statement

The authors have declared no competing interest.

### Author Declarations

Ethics committee of Peking University gave ethical approval for the China Health and Retirement Longitudinal Study (CHARLS). All participants provided written informed consent prior to participation. The present study is a secondary analysis of de-identified publicly available data.

## Reference

1. Livingston G, Huntley J, Liu KY, Costafreda SG, Selbæk G, Alladi S, et al. Dementia prevention, intervention, and care: 2024 report of the Lancet standing Commission. Lancet. 2024;404(10452):572–628.

2. McEwen BS, Morrison JH. The brain on stress: vulnerability and plasticity of the prefrontal cortex over the life course. Neuron. 2013;79(1):16–29.

3. Zhou Y, Kivimäki M, Holt-Lunstad J, Yan LL, Zhang Y, Wang H, et al. Stressful life events in childhood and adulthood and risk of physical, psychological and cognitive multimorbidities: a multicohort study. EClinicalMedicine. 2025;83:103225.

4. Li J, Ortí-Casañ N, Bayraktaroglu I, Mozzanica G, Zhang F, Olivier JDA, et al. Psychosocial stressors and cognitive function: An analysis using data from the English longitudinal study of ageing. The Journal of Prevention of Alzheimer’s Disease. 2025;12(8):100232.

5. Wijbenga L, Reijneveld SA, Almansa J, Korevaar EL, Hofstra J, de Winter AF. Trajectories of stressful life events and long-term changes in mental health outcomes, moderated by family functioning? the TRAILS study. Child and Adolescent Psychiatry and Mental Health. 2022;16(1):106.

6. Lupien SJ, McEwen BS, Gunnar MR, Heim C. Effects of stress throughout the lifespan on the brain, behaviour and cognition. Nat Rev Neurosci. 2009;10(6):434–45.

7. Popoli M, Yan Z, McEwen BS, Sanacora G. The stressed synapse: the impact of stress and glucocorticoids on glutamate transmission. Nat Rev Neurosci. 2011;13(1):22–37.

8. Yau JL, Seckl JR. Local amplification of glucocorticoids in the aging brain and impaired spatial memory. Front Aging Neurosci. 2012;4:24.

9. Justice NJ. The relationship between stress and Alzheimer’s disease. Neurobiol Stress. 2018;8:127–33.

10. Chen Y, Baram TZ. Toward Understanding How Early-Life Stress Reprograms Cognitive and Emotional Brain Networks. Neuropsychopharmacology. 2016;41(1):197–206.

11. McLaughlin KA, Sheridan MA, Lambert HK. Childhood adversity and neural development: deprivation and threat as distinct dimensions of early experience. Neurosci Biobehav Rev. 2014;47:578–91.

12. Lesuis SL, Hoeijmakers L, Korosi A, de Rooij SR, Swaab DF, Kessels HW, et al. Vulnerability and resilience to Alzheimer’s disease: early life conditions modulate neuropathology and determine cognitive reserve. Alzheimer’s Research & Therapy. 2018;10(1):95.

13. Hanson JL, Nacewicz BM, Sutterer MJ, Cayo AA, Schaefer SM, Rudolph KD, et al. Behavioral problems after early life stress: contributions of the hippocampus and amygdala. Biol Psychiatry. 2015;77(4):314–23.

14. Bisht K, Sharma K, Tremblay M. Chronic stress as a risk factor for Alzheimer’s disease: Roles of microglia-mediated synaptic remodeling, inflammation, and oxidative stress. Neurobiol Stress. 2018;9:9–21.

15. Bangasser DA, Valentino RJ. Sex differences in stress-related psychiatric disorders: neurobiological perspectives. Front Neuroendocrinol. 2014;35(3):303–19.

16. Levine DA, Gross AL, Briceño EM, Tilton N, Giordani BJ, Sussman JB, et al. Sex Differences in Cognitive Decline Among US Adults. JAMA Netw Open. 2021;4(2):e210169.

17. Stern Y. Cognitive reserve in ageing and Alzheimer’s disease. The Lancet Neurology. 2012;11(11):1006–12.

18. Meng X, D’Arcy C. Education and dementia in the context of the cognitive reserve hypothesis: a systematic review with meta-analyses and qualitative analyses. PLoS One. 2012;7(6):e38268.

19. Alesina A, Giuliano P, Nunn N. On the Origins of Gender Roles: Women and the Plough *. The Quarterly Journal of Economics. 2013;128(2):469–530.

20. Gneezy U, Leonard KL, List JA. Gender Differences in Competition: Evidence From a Matrilineal and a Patriarchal Society. Econometrica. 2009;77(5):1637–64.

21. Jayachandran S. The Roots of Gender Inequality in Developing Countries. Annual Review of Economics. 2015;7(Volume 7, 2015):63–88.

22. Zhao Y, Hu Y, Smith JP, Strauss J, Yang G. Cohort profile: the China Health and Retirement Longitudinal Study (CHARLS). Int J Epidemiol. 2014;43(1):61–8.

23. Sonnega A, Faul JD, Ofstedal MB, Langa KM, Phillips JW, Weir DR. Cohort Profile: the Health and Retirement Study (HRS). Int J Epidemiol. 2014;43(2):576–85.

24. Crimmins EM, Kim JK, Langa KM, Weir DR. Assessment of cognition using surveys and neuropsychological assessment: the Health and Retirement Study and the Aging, Demographics, and Memory Study. J Gerontol B Psychol Sci Soc Sci. 2011;66 Suppl 1(Suppl 1):i162–71.

25. Eskandari F, Salimi M, Binayi F, Abdollahifar MA, Eftekhary M, Hedayati M, et al. Investigating the Effects of Maternal Separation on Hypothalamic-Pituitary-Adrenal Axis and Glucose Homeostasis under Chronic Social Defeat Stress in Young Adult Male Rat Offspring. Neuroendocrinology. 2023;113(3):361–80.

26. Huang J, Shen C, Ye R, Shi Y, Li W. The Effect of Early Maternal Separation Combined With Adolescent Chronic Unpredictable Mild Stress on Behavior and Synaptic Plasticity in Adult Female Rats. Front Psychiatry. 2021;12:539299.

27. Sailer LL, Patel PP, Park AH, Moon J, Hanadari-Levy A, Ophir AG. Synergistic consequences of early-life social isolation and chronic stress impact coping and neural mechanisms underlying male prairie vole susceptibility and resilience. Front Behav Neurosci. 2022;16:931549.

28. McEwen BS. Protective and damaging effects of stress mediators. N Engl J Med. 1998;338(3):171–9.

29. Wade M, Zeanah CH, Fox NA, Tibu F, Ciolan LE, Nelson CA. Stress sensitization among severely neglected children and protection by social enrichment. Nature Communications. 2019;10(1):5771.

30. Gladish N, Merrill SM, Kobor MS. Childhood Trauma and Epigenetics: State of the Science and Future. Curr Environ Health Rep. 2022;9(4):661–72.

31. Bigio B, Sagi Y, Barnhill O, Dobbin J, El Shahawy O, de Angelis P, et al. Epigenetic embedding of childhood adversity: mitochondrial metabolism and neurobiology of stress-related CNS diseases. Front Mol Neurosci. 2023;16:1183184.

32. Bourassa KJ, Sbarra DA. Trauma, adversity, and biological aging: behavioral mechanisms relevant to treatment and theory. Translational Psychiatry. 2024;14(1):285.

33. Teicher MH, Samson JA, Anderson CM, Ohashi K. The effects of childhood maltreatment on brain structure, function and connectivity. Nat Rev Neurosci. 2016;17(10):652–66.

34. Andersen SL, Teicher MH. Stress, sensitive periods and maturational events in adolescent depression. Trends Neurosci. 2008;31(4):183–91.

35. McEwen BS, Nasca C, Gray JD. Stress Effects on Neuronal Structure: Hippocampus, Amygdala, and Prefrontal Cortex. Neuropsychopharmacology. 2016;41(1):3–23.

36. Kudielka BM, Kirschbaum C. Sex differences in HPA axis responses to stress: a review. Biological Psychology. 2005;69(1):113–32.

37. Christian LM, Wilson SJ, Madison AA, Prakash RS, Burd CE, Rosko AE, et al. Understanding the health effects of caregiving stress: New directions in molecular aging. Ageing Research Reviews. 2023;92:102096.

38. Subramaniapillai S, Almey A, Natasha Rajah M, Einstein G. Sex and gender differences in cognitive and brain reserve: Implications for Alzheimer’s disease in women. Front Neuroendocrinol. 2021;60:100879.

39. Xiong C, Biscardi M, Astell A, Nalder E, Cameron JI, Mihailidis A, et al. Sex and gender differences in caregiving burden experienced by family caregivers of persons with dementia: A systematic review. PLoS One. 2020;15(4):e0231848.

40. Pinquart M, Sörensen S. Gender Differences in Caregiver Stressors, Social Resources, and Health: An Updated Meta-Analysis. The Journals of Gerontology: Series B. 2006;61(1):P33–P45.

41. Hollanders JJ, van der Voorn B, Rotteveel J, Finken MJJ. Is HPA axis reactivity in childhood gender-specific? A systematic review. Biology of Sex Differences. 2017;8(1):23.

42. Barulli D, Stern Y. Efficiency, capacity, compensation, maintenance, plasticity: emerging concepts in cognitive reserve. Trends in Cognitive Sciences. 2013;17(10):502–9.

43. Raghupathi V, Raghupathi W. The influence of education on health: an empirical assessment of OECD countries for the period 1995–2015. Archives of Public Health. 2020;78(1):20.

44. Kang S, Lee J-L, Koo J-H. The buffering effect of social capital for daily mental stress in an unequal society: a lesson from Seoul. International Journal for Equity in Health. 2023;22(1):64.

45. Zajacova A, Lawrence EM. The Relationship Between Education and Health: Reducing Disparities Through a Contextual Approach. Annu Rev Public Health. 2018;39:273–89.

46. Glauber R. Rural depopulation and the rural-urban gap in cognitive functioning among older adults. J Rural Health. 2022;38(4):696–704.

47. Holt-Lunstad J. Social connection as a critical factor for mental and physical health: evidence, trends, challenges, and future implications. World Psychiatry. 2024;23(3):312–32.

48. Luo S, Chen W, Hu W, Wang HHX, Li J, Guo VY, et al. Parental Education, Own Education, and Cognitive Function in Middle-Aged and Older Adults. JAMA Netw Open. 2025;8(5):e2513036.

49. Zhang W, Chen Y, Chen N. Body mass index and trajectories of the cognition among Chinese middle and old-aged adults. BMC Geriatr. 2022;22(1):613.

