## Supplementary materials for "Life-course Stress Exposure and Cognitive Decline in Middle-aged and Older Chinese Adults: The Role of Sex Differences and Educational Protection"

**Supplementary Table 1.** Definition and assessment of main variables in CHARLS

| **Variables** | **Values** | **Measurements in CHARLS** |
| --- | --- | --- |
| **Childhood stressful life events** | | |
| Financial hardship | Yes | Before age 17, the R's self-rated family financial situation compared to the average family in the same community/village: 5. A lot worse off than them |
|  | No | Before age 17, the R's self-rated family financial situation compared to the average family in the same community/village: 1. A lot better off than them 2. Somewhat better off than them 3. Same as them 4. Somewhat worse off than them |
| Parental unemployment | Yes | Before age 17, the R's male or female guardian was ever unemployed and actively looked for a job. |
|  | No | Otherwise |
| Parental with substance abuse | Yes | During childhood, either of the R's guardians had an alcoholic nor had a drug problem. |
|  | No | Otherwise |
| Foster family/foster home | Yes | Before age 17, adopted mother or father spent the most time raising the R. |
|  | No | Otherwise |
| Parental separation or divorce | Yes | The R's age when parents divorced: 1. 0–3 years 2. 4–11 years 3. 12–16 years |
|  | No | The R's parents did not divorce; or the R's age when parents divorced: 1. 17 years old and older |
| Parental death | Yes | Before age 16, one or both R's biological parents died. |
|  | No | Otherwise |
| **Adulthood stressful life events** | | |
| Unemployment | Yes | The labor force status for the R: 6. unemployed |
|  | No | The labor force status for the R: 1. agricultural employed 2. agricultural self-employed 3. non-agricultural employed 4. non-agricultural self-employed 5. non-agricultural unpaid family business 7. retired 8. never worked. |
| Asset poverty | Yes | 0 or negative net worth of personal wealth |
|  | No | Positive net worth of personal wealth |
| Death of a child | Yes | Experience of the death of the R's own child |
|  | No | Otherwise |
| Death of a spouse/partner | Yes | Marital status: widowed |
|  | No | Marital status: Married, partnered, separated, divorced, and never married |
| Life-threatening illness or accident | Yes | Received medical treatment as the result of a traffic accident or another kind of major accidental injury |
|  | No | Otherwise |
| Physical attack/injury | Yes | Received a physical injury that has led to any permanent handicap, disability or limitations in what you can do in daily life |
|  | No | Otherwise |

**Note:** CHARLS, the China Health and Retirement Longitudinal Study; R, respondent.

**Supplementary Table 2.** Cross-sectional and longitudinal associations between stressful life events and memory score across adjustment models

| Model | None | Childhood only | Adulthood only | Childhood and adulthood |
| --- | --- | --- | --- | --- |
| Value | Ref | *β* (95% CI) | *β* (95% CI) | *β* (95% CI) |
| *Cross-sectional* |  |  |  |  |
| **Model 1** | Ref | -0.16 (-0.27, -0.05) | -0.07 (-0.18, 0.04) | -0.23 (-0.35, -0.10) |
|  |  | *p* <0.01^**^ | *p=*0.185 | *p* <0.001^***^ |
| **Model 2** | Ref | -0.10 (-0.21, -0.01) | -0.03 (-0.14, 0.07) | -0.14 (-0.27, -0.01) |
|  |  | *p* <0.05^*^ | *p*=0.529 | *p* <0.05^*^ |
| *Longitudinal* |  |  |  |  |
| **Model 1** | Ref | -0.26 (-0.34, -0.17) | -0.17 (-0.26, -0.08) | -0.37 (-0.47, -0.27) |
|  |  | *p* <0.001^***^ | *p* <0.001^***^ | *p* <0.001^***^ |
| **Model 2** | Ref | -0.18 (-0.26, -0.10) | -0.12 (-0.20, -0.04) | -0.26 (-0.35, -0.16) |
|  |  | *p* <0.001^***^ | *p* <0.01^**^ | *p* <0.001^***^ |

Notes: The memory score is calculated as the average of immediate and delayed recall, reflecting an individual's ability to remember recent events. Values represent regression coefficients (β), 95% confidence intervals, and p-values for the association between SLE exposure and cognitive function.
The reference group is individuals without any reported SLE exposure (“None”).
Model 1 was unadjusted.
Model 2 additionally adjusts for age, sex, education, residence, smoking and drinking status, BMI category, hypertension, diabetes, and cardiovascular disease.

^***^*p* < 0.001, ^**^*p* < 0.01, ^*^ *p* <0.05

**Supplementary Table 3.** Cross-sectional and longitudinal associations between stressful life events and executive score across adjustment models

| Model | None | Childhood only | Adulthood only | Childhood and adulthood |
| --- | --- | --- | --- | --- |
| Value | Ref | *β* (95% CI) | *β* (95% CI) | *β* (95% CI) |
| *Cross-sectional* |  |  |  |  |
| **Model 1** | Ref | -0.17 (-0.27, -0.08) | -0.24 (-0.34, -0.13) | -0.29 (-0.41, -0.17) |
|  |  | *p* <0.001^***^ | *p* <0.001^***^ | *p* <0.001^***^ |
| **Model 2** | Ref | -0.03 (-0.12, 0.05) | -0.10 (-0.22, -0.01) | -0.13 (-0.23, -0.04) |
|  |  | *p* =0.470 | *p* <0.05^*^ | *p* <0.01^**^ |
| *Longitudinal* |  |  |  |  |
| **Model 1** | Ref | -0.16 (-0.24, -0.08) | -0.19 (-0.27, -0.11) | -0.31 (-0.41, -0.21) |
|  |  | *p* <0.001^***^ | *p* <0.001^***^ | *p* <0.001^***^ |
| **Model 2** | Ref | -0.02 (-0.08, 0.05) | -0.07 (-0.14, -0.01) | -0.11 (-0.19, -0.02) |
|  |  | *p* =0.662 | *p* <0.05^*^ | *p* <0.05^*^ |

Notes: The executive score is derived from serial subtraction by sevens (5 times) and a figure-drawing task, reflecting the respondent’s executive functions. Values represent regression coefficients (β), 95% confidence intervals, and p-values for the association between SLE exposure and cognitive function.
The reference group is individuals without any reported SLE exposure (“None”).
Model 1 was unadjusted.
Model 2 additionally adjusts for age, sex, education, residence, smoking and drinking status, BMI category, hypertension, diabetes, and cardiovascular disease.

^***^*p* < 0.001, ^**^*p* < 0.01, ^*^ *p* <0.05

**Supplementary Table 4.** Cross-sectional and longitudinal associations between stressful life events and orientation score across adjustment models

| Model | None | Childhood only | Adulthood only | Childhood and adulthood |
| --- | --- | --- | --- | --- |
| Value | Ref | *β* (95% CI) | *β* (95% CI) | *β* (95% CI) |
| *Cross-sectional* |  |  |  |  |
| **Model 1** | Ref | -0.09 (-0.15, -0.04) | -0.10 (-0.16, -0.05) | -0.20 (-0.27, -0.13) |
|  |  | *p* <0.01^**^ | *p* <0.001^***^ | *p* <0.001^***^ |
| **Model 2** | Ref | -0.01 (-0.06, 0.05) | -0.05 (-0.10, 0.01) | -0.08 (-0.15, -0.01) |
|  |  | *p* =0.851 | *p* =0.124 | *p* <0.05^*^ |
| *Longitudinal* |  |  |  |  |
| **Model 1** | Ref | -0.12 (-0.16, -0.08) | -0.13 (-0.17, -0.09) | -0.20 (-0.25, -0.15) |
|  |  | *p* <0.001^***^ | *p* <0.001^***^ | *p* <0.001^***^ |
| **Model 2** | Ref | -0.02 (-0.06, 0.02) | -0.07 (-0.11, -0.03) | -0.08 (-0.12, -0.03) |
|  |  | *p* =0.331 | *p* <0.01^**^ | *p* <0.01^**^ |

Notes: The orientation score is based on the correctness of responses to the current year, month, date, day of week and season, reflecting the respondent’s awareness of temporal information. Values represent regression coefficients (β), 95% confidence intervals, and p-values for the association between SLE exposure and cognitive function.
The reference group is individuals without any reported SLE exposure (“None”).
Model 1 was unadjusted.
Model 2 additionally adjusts for age, sex, education, residence, smoking and drinking status, BMI category, hypertension, diabetes, and cardiovascular disease.

^***^*p* < 0.001, ^**^*p* < 0.01, ^*^ *p* <0.05

**Supplementary Table 5.** Interaction effects of age, sex, and education with SLE exposure on global cognitive function

|  | Age × Group | | Sex × Group | | Education × Group | |
| --- | --- | --- | --- | --- | --- | --- |
| *Predictors* | *β* (95% CI) | *p* | *β* (95% CI) | *p* | *β* (95% CI) | *p* |
| **SLEs** (Ref: None) |  |  |  |  |  |  |
| Childhood only | -0.13 (-0.41, 0.16) | 0.373 | -0.21 (-0.36, -0.05) | <0.01**^**^** | -0.15 (-0.34, 0.02) | 0.104 |
| Adulthood only | -0.02 (-0.25, 0.24) | 0.980 | -0.13 (-0.28, 0.02) | 0.098 | -0.11 (-0.29, 0.08) | 0.243 |
| Childhood and adulthood | -0.14 (-0.37, 0.10) | 0.261 | -0.25 (-0.43, -0.07) | <0.01**^**^** | -0.42 (-0.63, -0.19) | <0.001**^***^** |

Notes: Interaction effects between SLE exposure and age, sex, and education on global cognitive function. Coefficients (β) are shown with 95% confidence intervals and p-values. All interactions were evaluated using bootstrap resampling (R = 1000).

Reference categories: No SLE exposure, younger age, male, high education.

^***^ *p* < 0.001, ^**^*p* < 0.01, ^*^ *p* <0.05

**Supplementary Table 6.** Sex-stratified associations between stressful life events and global cognitive function across adjustment models

| Model | None | Childhood only | Adulthood only | Childhood and adulthood |
| --- | --- | --- | --- | --- |
| Value | Ref | *β* (95% CI) | *β* (95% CI) | *β* (95% CI) |
| *Male* |  |  |  |  |
| **Model 1** | Ref | -0.48 (-0.66, -0.30) | -0.39 (-0.58, -0.19) | -0.83 (-1.04, -0.62) |
|  |  | *p* <0.001^***^ | *p* <0.001^***^ | *p* <0.001^***^ |
| **Model 2** | Ref | -0.44 (-0.62, -0.26) | -0.38 (-0.57, -0.20) | -0.79 (-1.00, -0.58) |
|  |  | *p* <0.001^***^ | *p* <0.001^***^ | *p* <0.001^***^ |
| *Female* |  |  |  |  |
| **Model 1** | Ref | -0.70 (-0.96, -0.44) | -0.59 (-0.85, -0.34) | -1.14 (-1.46, -0.83) |
|  |  | *p* <0.001^***^ | *p* <0.001^***^ | *p* <0.001^***^ |
| **Model 2** | Ref | -0.65 (-0.90, -0.39) | -0.51 (-0.76, -0.26) | -1.02 (-1.33, -0.71) |
|  |  | *p* <0.001^***^ | *p* <0.001^***^ | *p* <0.001^***^ |

Notes: Values represent regression coefficients (β), 95% confidence intervals, and p-values for the association between SLE exposure and global cognitive function.
The reference group is individuals without any reported SLE exposure (“None”).
Model 1 was unadjusted.
Model 2 additionally adjusts for age, education, residence, smoking and drinking status, BMI category, hypertension, diabetes, and cardiovascular disease.

^***^*p* < 0.001, ^**^*p* < 0.01, ^*^ *p* <0.05

**Supplementary Table 7.** Education-stratified associations between stressful life events and global cognitive function across adjustment models

| Model | None | Childhood only | Adulthood only | Childhood and adulthood |
| --- | --- | --- | --- | --- |
| Value | Ref | *β* (95% CI) | *β* (95% CI) | *β* (95% CI) |
| *Below high school* |  |  |  |  |
| **Model 1** | Ref | -0.35 (-0.54, -0.15) | -0.29 (-0.48, -0.09) | -0.79 (-0.98, -0.59) |
|  |  | *p* <0.001^***^ | *p* <0.01^**^ | *p* <0.001^***^ |
| **Model 2** | Ref | -0.30 (-0.49, -0.10) | -0.20 (-0.39, -0.01) | -0.57 (-0.76, -0.37) |
|  |  | *p* <0.01^**^ | *p* <0.05^*^ | *p* <0.001^***^ |
| *High school or above* |  |  |  |  |
| **Model 1** | Ref | -0.33 (-0.66, -0.01) | -0.28 (-0.73, 0.16) | -0.40 (-0.66, -0.14) |
|  |  | *p* <0.05^*^ | *p* =0.210 | *p* <0.01^**^ |
| **Model 2** | Ref | -0.16 (-0.41, 0.10) | -0.13 (-0.49, 0.22) | -0.27 (-0.52, -0.01) |
|  |  | *p* =0.233 | *p* =0.455 | *p* <0.05^*^ |

Notes: Values represent regression coefficients (β), 95% confidence intervals, and p-values for the association between SLE exposure and cognitive function.
The reference group is individuals without any reported SLE exposure (“None”).
Model 1 was unadjusted.
Model 2 additionally adjusts for age, sex, residence, smoking and drinking status, BMI category, hypertension, diabetes, and cardiovascular disease.

^***^*p* < 0.001, ^**^*p* < 0.01, ^*^ *p* <0.05
